# Risk factors for severe complications and mortality among hospitalized children with measles during a major outbreak in Northern Vietnam, 2017 - 2019

**DOI:** 10.1101/2023.07.16.23292745

**Authors:** Minh Dien Tran, Nhung T. H. Pham, Hoang Nguyen Vu, Minh-Hung Tran, Hoang-Anh Ngo, Phuc H Phan

## Abstract

**Introduction:** Measles outbreaks increased worldwide during the period of 2017-2019. Similarly, Vietnam experienced one of the largest measles outbreaks in recent decades, with various paediatric patients presenting severe complications. In this study, we aim to identify factors associated with death and severity among children with measles admitted to Vietnam National Children’s Hospital (VNCH) in Hanoi, Vietnam between 2017 and 2019.

**Method:** This single-center retrospective cohort study included 2, 072 patients with measles admitted to VNCH from 1/1/2017 to 31/12/2019. Data on epidemiological, clinical characteristics, vaccine status, and outcomes were collected and summarised. We conducted both univariable and multivariable logistic regression analyses to examine the correlations between various characteristics of hospitalized children and mortality.

**Findings:** In total, there were 2, 072 patients, including 1, 297 (62.6%) males and 775 (37.4%) females. The median age was 9 months (interquartile range 7 *−* 17). 87.3% of cases had not received any measles-containing vaccine (MCV). 30 (1.4%) patients died, with 40% aged less than 9 months. Only 3 among 30 (10%) who died had received at least 1 dose of MCV. Bronchopneumonia was the most common complication, occurring in 1, 413 (68.2%) patients. The following characteristics were significantly associated with mortality in the multivariable analysis: age under 9 months and age from 9 months to 5 years, residing 20 to 200 kilometres from VNCH, and having co-infection with adenovirus or other hospital acquired infections. Age group was also significantly associated with severity in the multivariable analysis.

**Interpretation:** Vietnam continues to face the threat of future measles epidemics, given the burden of hospitalization and the high rate of complications observed in hospitalized patients. This highlights the critical need to maintain high measles vaccine coverage, particularly by targeting the unvaccinated population. To prevent future outbreaks and lower measles incidence, routine immunization needs to be strengthened, and earlier scheduling of MCV1 needs to be further evaluated. The comprehensive analysis of the 2017-19 measles outbreak presented in this study will contribute to informed decision-making regarding appropriate measures to counteract future resurgences of measles in Vietnam.

**Funding:** No specific grant from funding agencies in the public, commercial or not-for-profit sectors supported the submission and publication of this manuscript.

## 1. Introduction

Measles is considered one of the most contagious diseases for humans with the basic reproduction number of 12 *−* 18, which means that each person with measles would, on average, infects 12 *−* 18 other people in a totally susceptible population [1]. This disease is caused by a paramyxovirus virus, manifesting as a febrile rash illness. It is an airborne disease, which can easily spread through susceptible populations, and the only effective measure for preventing this disease is vaccination. Before the invention and widespread use of the measles vaccine in 1963, frequent outbreaks of the disease occurred approximately every 2–3 years, resulting in approximately 2.6 million deaths annually [2]. It was estimated that measles infected over 90% of children before they reached 15, causing over two million deaths and 15,000 to 60,000 cases of blindness worldwide each year [3, 4]. Accelerated immunization activities have significantly reduced the number of measles deaths. From 2000 to 2018, measles vaccination is estimated to have prevented 23.2 million deaths, resulting in a 73% decrease in global measles deaths from 536, 000 in 2000 to 132, 490 in 2016 [3, 5, 6]. Measles outbreaks happen when people who are not immune to the measles virus are infected and spread the disease to unvaccinated or under-vaccinated populations. To control measles and avoid outbreaks and deaths, at both national and sub-national levels, vaccination coverage rates must reach and be maintained at 95% for both the first and second doses. However, global vaccination coverage rates for first-dose measles-containing vaccine (MCV1) have stagnated at 84-85% for over a decade, and while second-dose measles-containing vaccine (MCV2) coverage rates have been gradually increasing, they are only at 71%. Measles vaccination rates remain significantly below the necessary 95% threshold to control measles and prevent outbreaks [5].

From 2017, measles experienced a surge worldwide, resulting in the highest number of reported cases in 23 years in 2019 [8]. World Health Organization (WHO) and the United States Centers for Disease Control and Prevention (CDC) published a report indicating that the number of reported measles cases increased to 869,770 in 2019, the highest number recorded since 1996, with all WHO regions reporting increases. The number of reporting countries with annual measles incidence of less than 5 cases per 1 million population decreased from 125 in 2016 to 85 in 2019 and an estimated 207,500 people worldwide died from measles, a nearly 50% increase since 2016 [5]. Measles cases surged drastically in WHO’s African Region (AFR) by 1, 606%, in the Region of the Americas (AMR) by 19, 739%, and in the European Region (EUR) by 2, 282% since 2016. 9 countries (Central African Republic, Democratic Republic of the Congo, Georgia, Kazakhstan, Madagascar, North Macedonia, Samoa, Tonga, and Ukraine), accounting for 73% of all reported cases worldwide, experienced large outbreaks with reported measles incidence exceeding 500 per 1 million population [6].

Similar to other countries, between 2017 and 2019, Vietnam experienced one of the largest measles outbreaks in recent decades. In 2018, Vietnam had 8,444 cases of typhus-suspected measles with 1,177 confirmed measles cases, twice as many cases as in 2017. Most measles cases were unvaccinated or incompletely vaccinated children, or had no known vaccination history due to lost immunization records. To prevent the outbreak, the Ministry of Health of Vietnam launched an additional measles-rubella vaccination campaign in late 2018 for 4.2 million children aged between 1 and 5 in vulnerable areas in 57 cities and provinces [9]. In 2019, Vietnam reported over 39,000 suspected cases of typhus linked to measles, with more than 7,100 confirmed cases and four deaths [9, 10]. Analysis of the total infections in Hanoi until April 2019 revealed that up to 14% of children under 9 months had measles. This is the group of children who have not yet reached the age of vaccination and the mother has not been vaccinated against measles, so they do not receive any antibodies during pregnancy. Children between 9 months and 5 years old had the highest incidence of the disease, accounting for 42% of total cases. Furthermore, the Center for Preventive Medicine has reported that approximately 15% of children of vaccination age are not received the required two full doses of the measles vaccine [11]. The objective of this study is to analyze epidemiological and clinical information of over 2,000 children with measles admitted to Vietnam National Children’s Hospital (VNCH), the national referral hospital for children in Hanoi, the capital city of Vietnam, between 1 January 2017 and 31 December 2019. We aim to provide an overview of the outbreak in the northern region of Vietnam and assess the potential implications for Vietnam and neighbouring countries.

## 2. Method

### 2.1. Study design and participants

A single-center retrospective cohort study including all children admitted with measles admitted to VNCH between 01/01/2017 and 31/12/2019 was undertaken. During this time, individuals with suspected measles were admitted to VNCH, in addition to other hospitals in the region. Admitted children included either self-referrals (walk-in patients) or referrals from other health facilities. Patients with multiple missing data were excluded.

### 2.2. Ethics statement

The study was approved by the Ethics Committee for Biomedical Research Vietnam National Children’s Hospital (Reference number: 473/BVNTW-HDDD, IRB identifier: IRB-VN01037/IRB00011976/FWA00028418). As the study involved a retrospective analysis of routinely collected data under strict anonymity, patient or caregiver consent was waived.

### 2.3. Procedures

Following WHO-recommended surveillance standard, the case definition of measles infection was based on laboratory-confirmed diagnosis. Eligible study participants are those who were diagnosed with measles by IgM or PCR test at any time point during their hospital stay, and vaccine-associated illness has been ruled out. All individuals from VNCH fulfilling the clinical case definition acquired information using the measles suspected case inquiry form. The information in the questionnaire was collected by interviewing parents about vaccination status in children’s vaccination records and using paper or electronic medical records to collect other information. Duplicates and unidentified patients are detected and removed by comparing the patient IDs. The date of admission, measles vaccination status according to the vaccination card (if available), diagnosis upon admission, and location of exposure according to the caregiver report, as well as socio-demographic, clinical features, and outcome, were all included. There was no information provided about the rash’s features, dietary status, or whether the case was likely a main or secondary home case.

Furthermore, to facilitate the analysis of factors associated with the risk of severe diseases, we defined severe patients as those requiring any form of oxygen supplementation and respiratory support, namely oxygen supplementation via nasal cannula, continuous positive airway pressure (CPAP), conventional mechanical ventilation, high-frequency oscillatory (HFO) ventilation, or extracorporeal membrane oxygenation (ECMO). To build an anonymised dataset, selected data were extracted and personal information such as name, date of birth, and permanent address were removed.

### 2.4. Statistical analysis

Univariable and multivariable Firth’s Bias-Reduced logistic regression analysis was used to assess associations between specific characteristics of hospitalized children and clinical outcomes (survived or died). Comparisons were made between three separate locations of exposure, and the Chi-squared or Fisher’s exact test was employed for the comparison of dichotomous and t-test for continuous variables. Odds ratios (ORs) were computed with a 95% confidence interval (CI), and a p-value less than or equal to 0.05 was considered statistically significant.

The selection of variables for the multivariable analysis was based on several factors. Basic variables such as year of hospitalization, vaccination status (patient having at least one dose of MCV), age, and exposure location were included as they are fundamental factors to consider. Additionally, the distance from the hospital was incorporated as it is expected to have an influence on the time taken to reach the hospital and the duration between symptom onset and testing/admission. Patients residing far from VNCH may initially receive treatment at lower-level hospitals, leading to potential delays in obtaining the correct treatment and adequate level of care. In addition to the previously mentioned factors, the duration of stay in the hospital was included as a variable in the analysis. This choice was motivated by the understanding that the duration of hospitalization can serve as an indicator of the severity of measles. Prolonged hospital stays may suggest a more severe manifestation of the disease. Co-infections and underlying conditions of several bodily systems were also added after ensuring that their correlation with other model covariates would not confound the link between the other variables and death. In the model analysing the risk towards severe diseases, the duration between symptom onset and testing was included as a variable based on its statistical significance. Conversely, in the mortality model, the variables “RSV” (respiratory syncytial virus) and “pertussis” (within the co-infection group of variables) were excluded due to their lack of statistical significance. We report the median and interquartile range (IQR) for age groups and the frequency and percentage for categorical variables. Despite the huge number of univariable tests, we did not explicitly apply a multiple-testing correction since the variable selection technique for the multivariable model was only partially based on p-values. The Pandas software library was used to arrange the data, and the R package “logistf” was utilized for the main analysis [12, 13]. For mapping and distance computation from patient’s place of residence to the hospital, the Azure Maps Route and Geopy libraries were used [14, 15].

All conducted statistical analysis can be found within the following GitHub repository.

## 3. Results

Within three years from 1/1/17 to 31/12/19, VNCH admitted 2,072 children who met the clinical case definition for measles.

Figure 2 and 3 depict the residence of hospitalized children throughout the study period, the number of measles cases admitted to VNCH each week, and the number of deceased and critical children. The measles outbreak in Vietnam surged in 2018-19, mirroring the trend of VNCH admissions, which increased from low levels in 2017 to over 50 cases per 2 weeks (25 cases per week) during the months of April and May 2019, before declining to pre-epidemic levels by September 2019. Figure 5 illustrates number of measles cases admitted to VNCH and total measles cases in Vietnam, 2017-2019. From 2017 to 2019, VNCH admitted 132, 621, and 1,319 measles cases, accounting for 58%, 27% and 9% of total measles cases in Vietnam.

**Figure 1:**
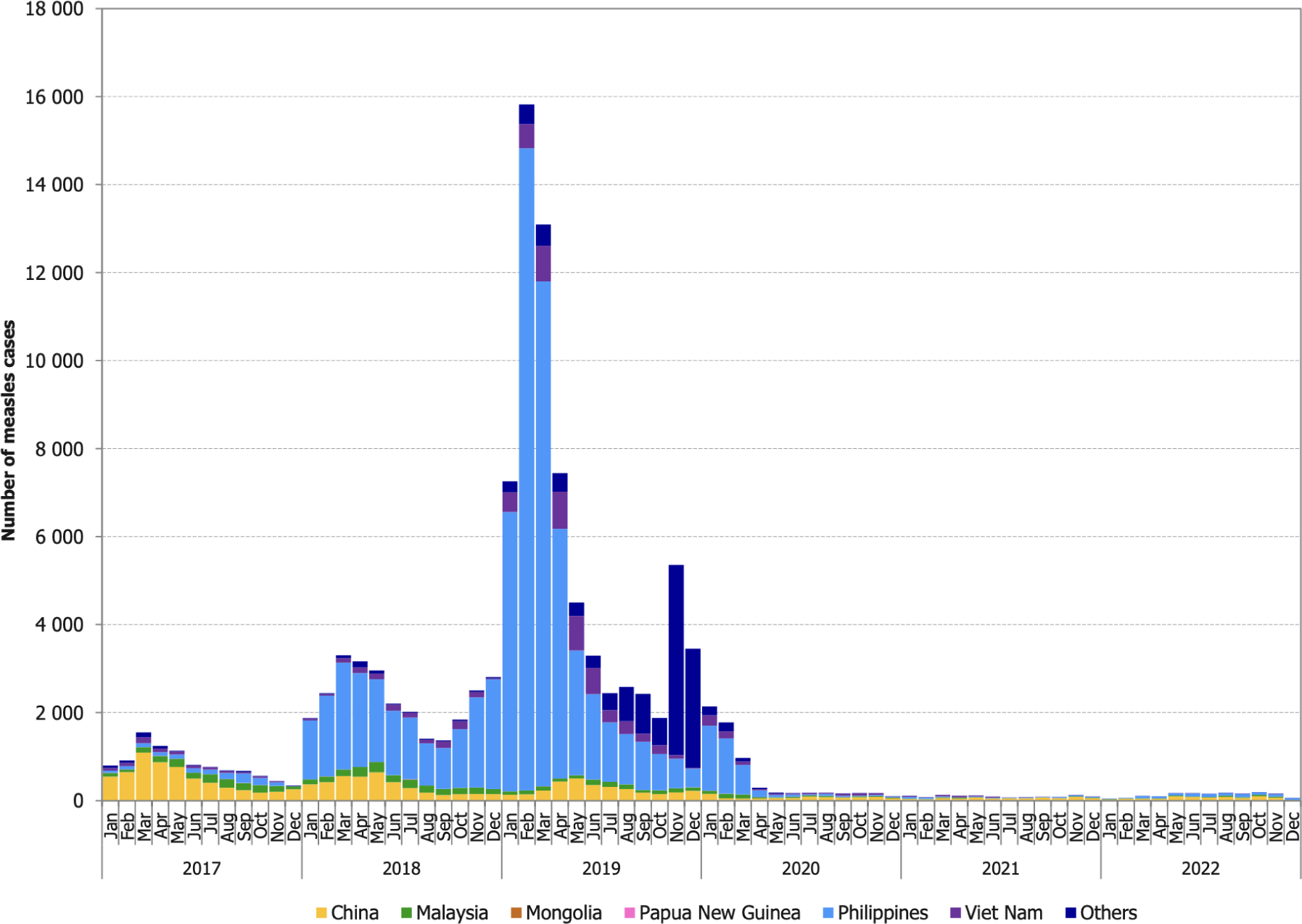
Time-series of reported confirmed and compatible measles cases in the Western Pacific Region by month of rash onset, 2017-2022 (adapted from: WHO Measles-Rubella Bulletin [7])

**Figure 2:**
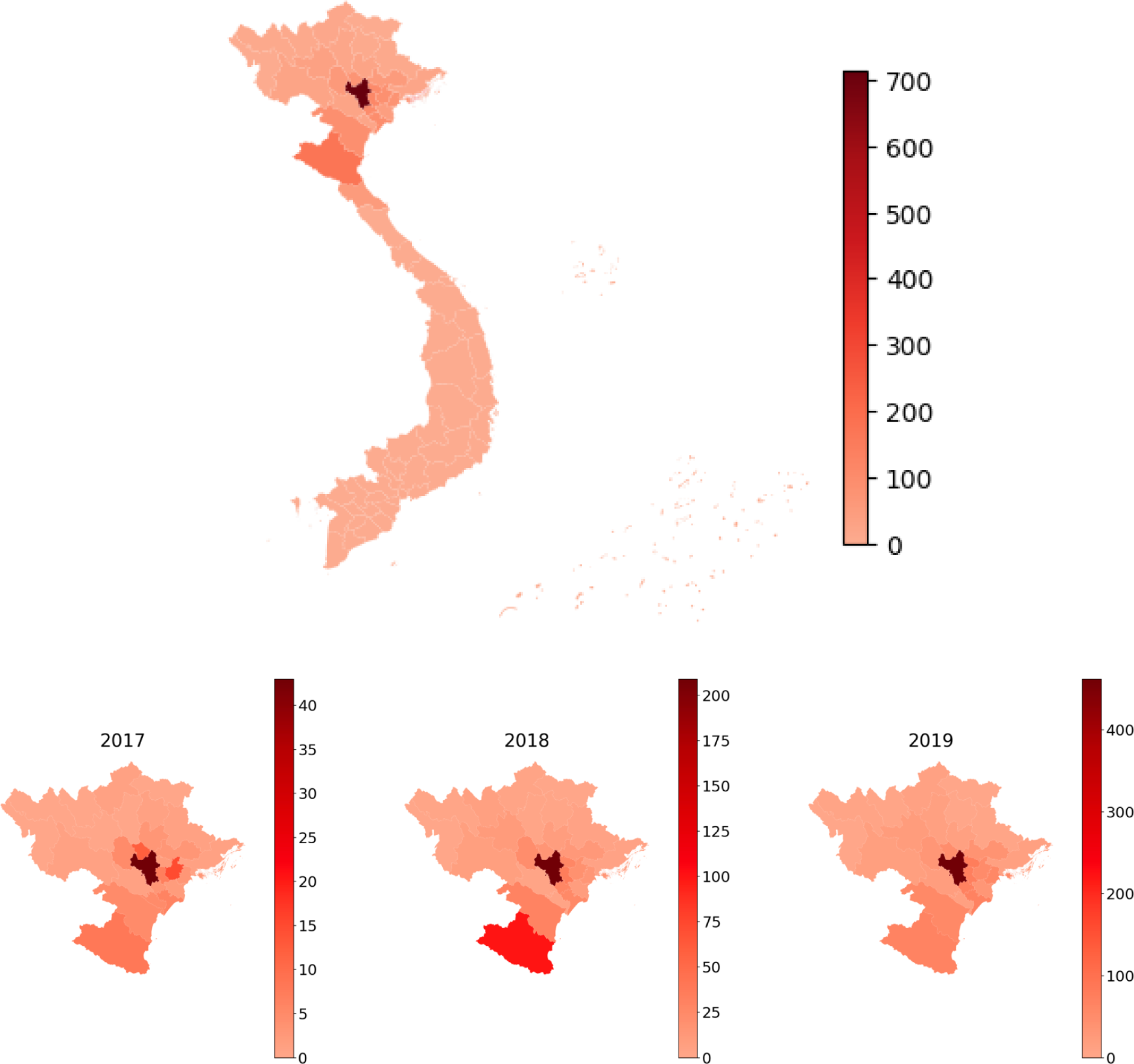
Distribution of hospitalized measles patients admitted to VNCH by provinces and city, and in northern Vietnam, 2017-2019.

**Figure 3:**
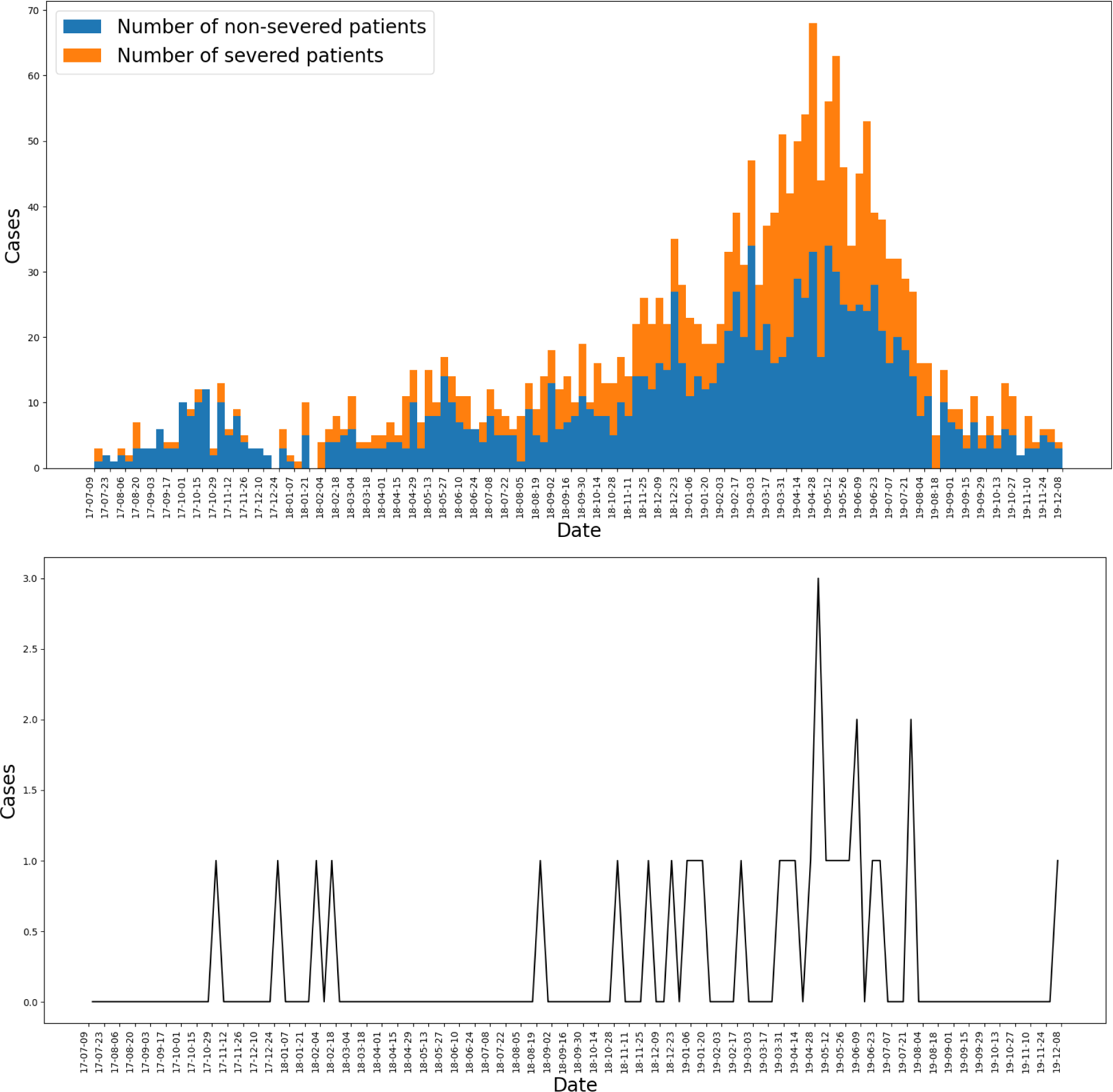
Epidemic curve of the hospitalized measles cases at VNCH by week of admission.

**Figure 4:**
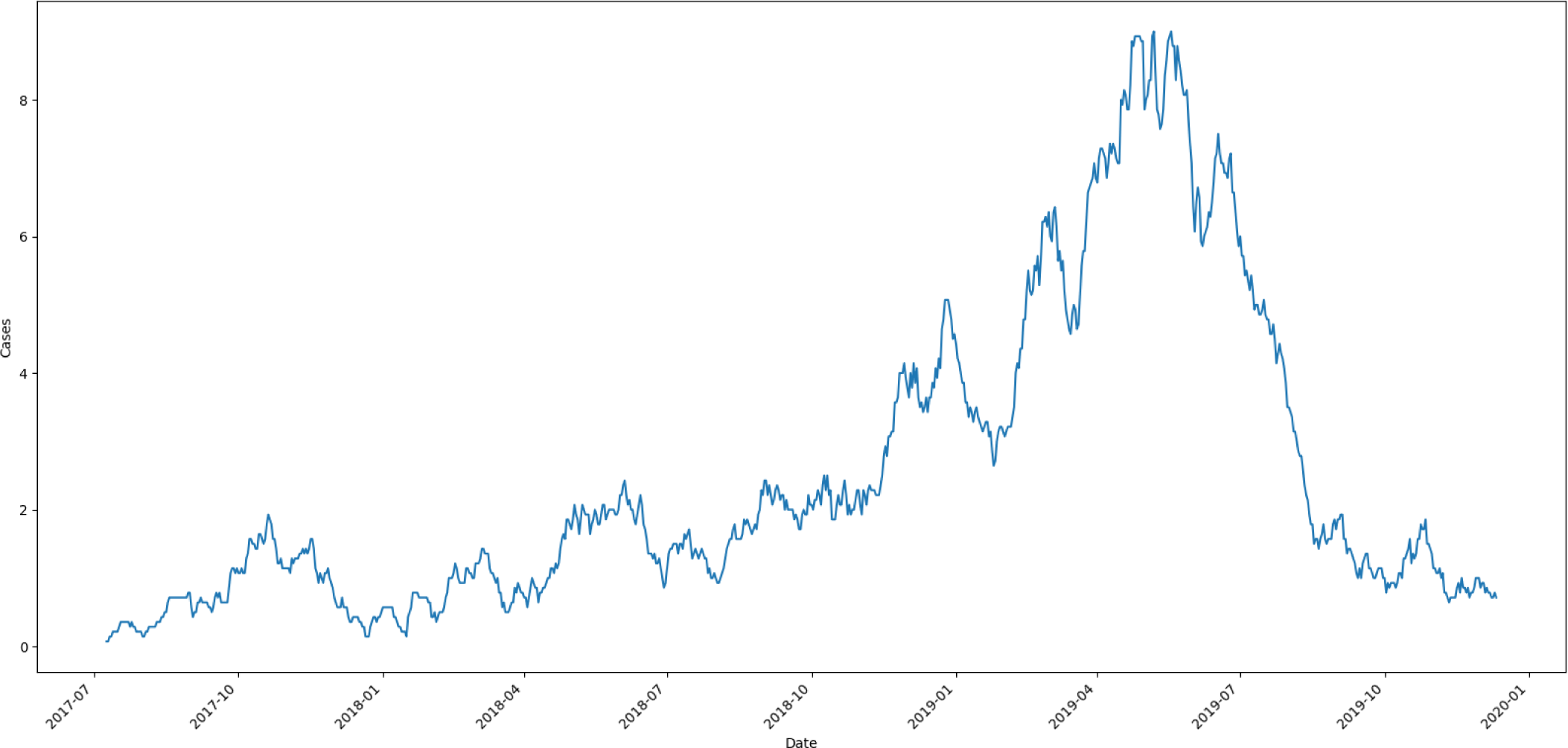
Daily trends in number of measles cases admitted to VNCH, with 7-day moving average.

**Figure 5:**
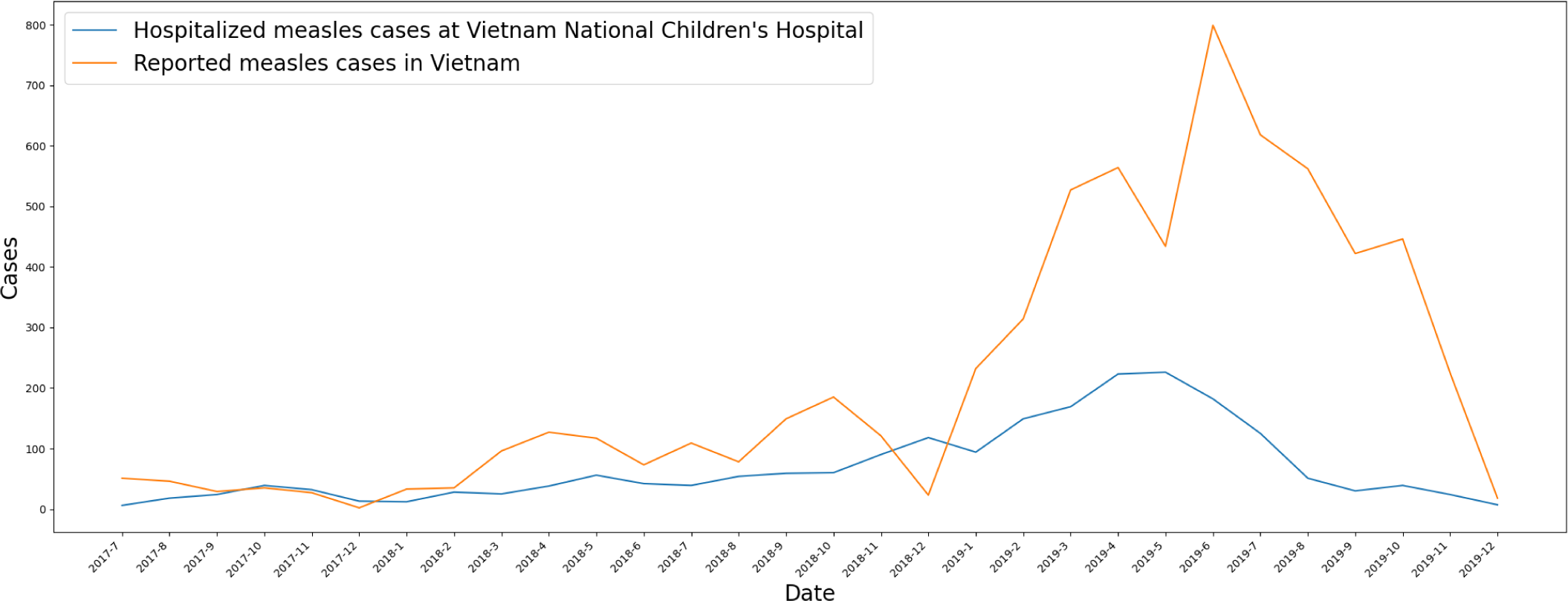
Hospitalized measles cases at Vietnam National Children’s Hospital and reported measles cases in Vietnam by months, 2017-2019.

The demographic and clinical features of the 2,072 children hospitalized with measles are described in Table 1. In 2017, there were 132 admissions and it quintuple in 2018 with 621 measles cases; more than half of the cases are in 2019 with 1,319 (63.7%) cases. 1,297 (62.6%) hospitalized children were male and 775 (37.4%) were female. The median age was 9.6 months, with an interquartile range (IQR) of 7 to 16.9 months; 43.5% were younger than 9 months, 47.5% were aged 9 months to 5 years, and 9% were older than 5 years. Most children (87.3%) were reported not to have received any MCV and only 9.8% received at least one dose. Measles cases were transmitted from the community in 61.8% of cases, from VNCH in 26.4% of cases, and from other hospitals in 11.6% of cases. 69.3% of patients live in the Red River Delta, including Hanoi, and 84.6% live within 200 kilometers of the hospital. In 66% of all instances, the period between the development of the rash and hospital admission was shorter than one week. Furthermore, 611 (29.5%) cases were admitted to the hospital before the onset date due to a different disease. More than half of patients (50.3%) are hospitalized for less than 1 week and 86% for less than three weeks. Most children (76.5%) have no underlying diseases, with only 4.5% having chronic diseases related to gastrointestinal system and 3.5% having chronic diseases of the neurological system. 849 (41%) patients turned severe after hospitialization, with 685 (33.1%) requiring oxygen supplementation via nasal cannula, and 144 patients requiring conventional mechanical ventilation. Complications were reported in most children, the most prevalent of which was bronchopneumonia (68.2%). Middle ear infection and febrile seizure complications were reported in 30 (1.4%) and 10 (0.5%) children, respectively.

**Table 1:** Demographic and clinical characteristics of hospitalized children with measles.

| **Characteristics ($N = 2,072$)** | **$N(\%)$ or median (IQR)** |
| --- | --- |
| Year of admission |  |
| 2017 | 132 (6.4) |
| 2018 | 621 (30.0) |
| 2019 | 1,319 (63.7) |
| Sex |  |
| Male | 1,297 (62.6) |
| Female | 775 (37.4) |
| Age in months (median, IQR) | 9.6 (7.0, 16.9) |
| Age group (in months) |  |
| 0 – 9 | 902 (43.5) |
| 9 – 60 | 984 (47.5) |
| $\geq 60$ | 186 (9.0) |
| Vaccination status |  |
| Unvaccinated | 1,808 (87.3) |
| Vaccinated (any dose) | 264 (12.7) |
| Place of exposure |  |
| Community | 1,281 (61.8) |
| Healthcare facility (including VNCH and other) | 788 (38.0) |
| Unknown | 3 (0.1) |
| Distance from the hospital (km) |  |
| 0 – 20 | 468 (22.6) |
| 20 – 200 | 1285 (62.0) |
| 200 – 500 | 305 (14.7) |
| $\geq 500$ | 14 (0.7) |
| Region of residence |  |
| Hanoi | 714 (34.5) |
| Northeastern region | 100 (4.8) |
| Northwestern region | 207 (10.0) |
| Red River Delta (except Hanoi) | 722 (34.8) |
| Central region | 321 (15.5) |
| Southern region | 8 (0.4) |
| Clinical outcome |  |
| Deceased | 30 (1.4) |
| Survived | 2,042 (98.6) |
| Duration between onset and admission |  |
| Admitted before the onset date due to a different disease ($\leq 0$) | 611 (29.5) |
| 0 – 3 | 916 (44.2) |
| 3 – 7 | 452 (21.8) |
| 7 – 14 | 47 (2.3) |
| $\geq 14$ | 46 (2.2) |
| Duration of stay within the hospital |  |
| 0 – 7 | 1,042 (50.3) |
| 7 – 21 | 739 (35.7) |
| $\geq 21$ | 291 (14.0) |
| Underlying conditions |  |
| Respiratory system | 42 (2.0) |
| Cardiovascular system | 72 (3.5) |
| Gastrointestinal system | 94 (4.5) |
| Kidney and urology system | 46 (2.2) |
| Immunodeficiency | 12 (0.6) |
| Neurological system | 73 (3.5) |
| Inherited metabolic disorders | 15 (0.7) |
| Other underlying conditions | 159 (7.7) |
| No underlying diseases | 1,585 (76.5) |
| Maximal form of respiratory support used |  |
| Oxygen supplementation via nasal cannula | 685 (33.1) |
| Continuous Positive Airway Pressure (CPAP) | 8 (0.4) |
| Conventional mechanical ventilation | 144 (0.9) |
| High-frequency Oscillatory (HFO) Ventilation | 11 (0.5) |
| Extracorporeal membrane oxygenation (ECMO) | 1 (0.0) |
| Duration between onset and test (detection time) (in hours) |  |
| Tested before onset ($<0$) | 23 (1.1) |
| 0 – 24 | 554 (26.7) |
| 24 – 48 | 438 (21.1) |
| $\geq 48$ | 1,057 (51.0) |
| Diagnosis upon admission |  |
| Measles | 848 (40.9) |
| Bronchopneumonia | 849 (41.0) |
| Other diagnosis | 391 (18.9) |
| Complications |  |
| Gastroenteritis | 51 (2.5) |
| Middle ear infection | 30 (1.4) |
| Conjunctivitis | 2 (0.1) |
| Laryngitis | 7 (0.3) |
| Pneumonia bronchitis | 1,413 (68.2) |
| Febrile seizures | 10 (0.5) |
| Sepsis/Septic shock | 4 (0.2) |
| Co-infections |  |
| Influenza A | 16 (0.8) |
| Influenza B | 4 (0.2) |
| *Staphylococcus aureus* | 3 (0.1) |
| *Streptococcus pneumonia* | 2 (0.1) |
| Respiratory Syncytial Virus (RSV) | 30 (1.4) |
| Adenovirus | 95 (4.6) |
| Pertussis | 17 (0.8) |
| Other co-infections | 18 (0.9) |
| Clinical classification |  |
| Severe | 849 (41.0) |
| Not severe | 1,223 (59.0) |

Overall, 30 (1.4%) children died and 2,041 (98.6%) were discharged.

Table 2 shows the demographic and clinical characteristics of hospitalized patients with measles by place of exposure, including community, with VNCH or from another health care facilities. These characteristics were significantly associated with exposure location: admission year, age groups, vaccination status, region of residence, clinical outcome, underlying conditions, maximal form of respiratory used, diagnosis upon admission, different types of coinfections, and clinical progression.

**Table 2:**
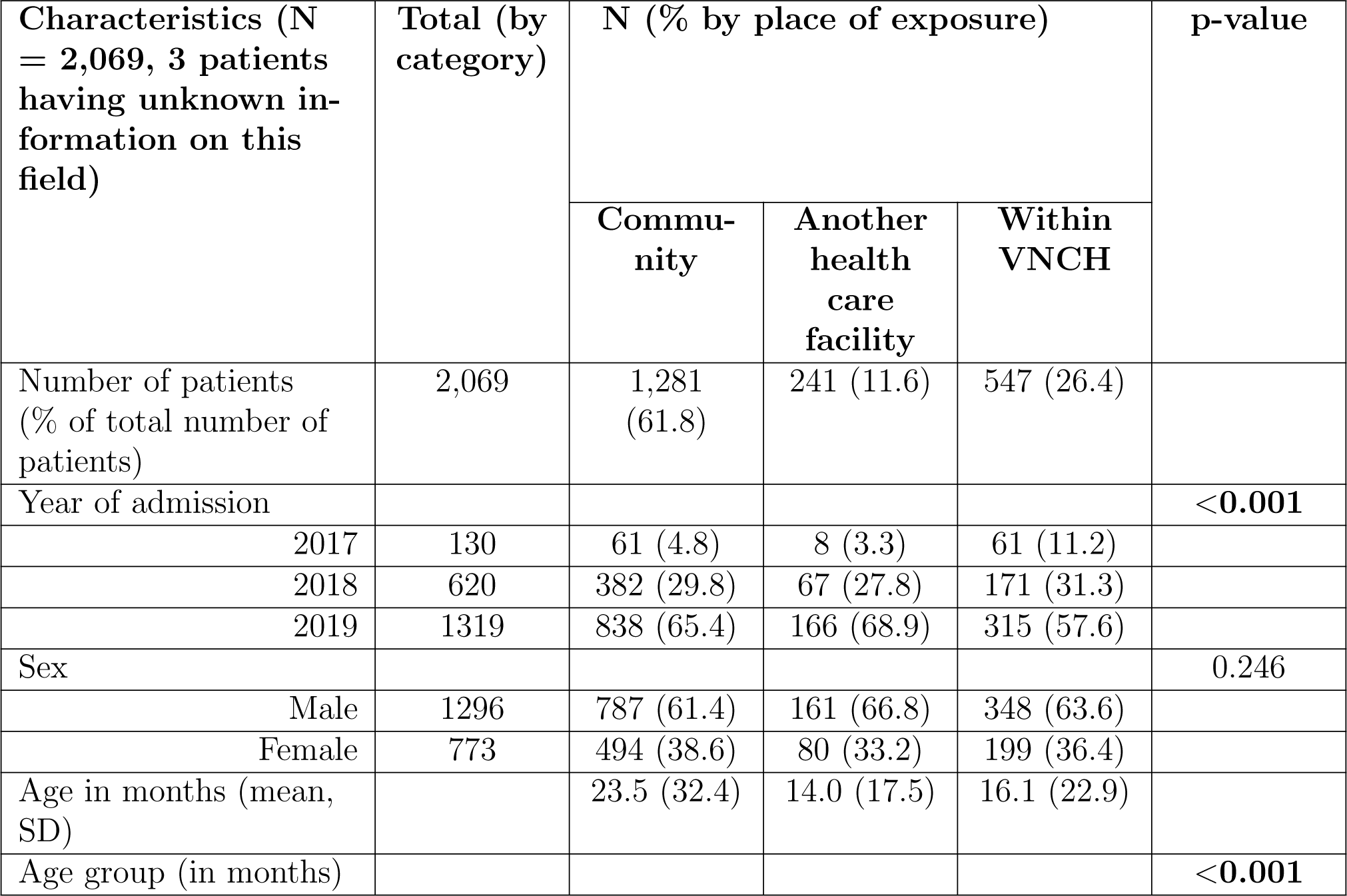
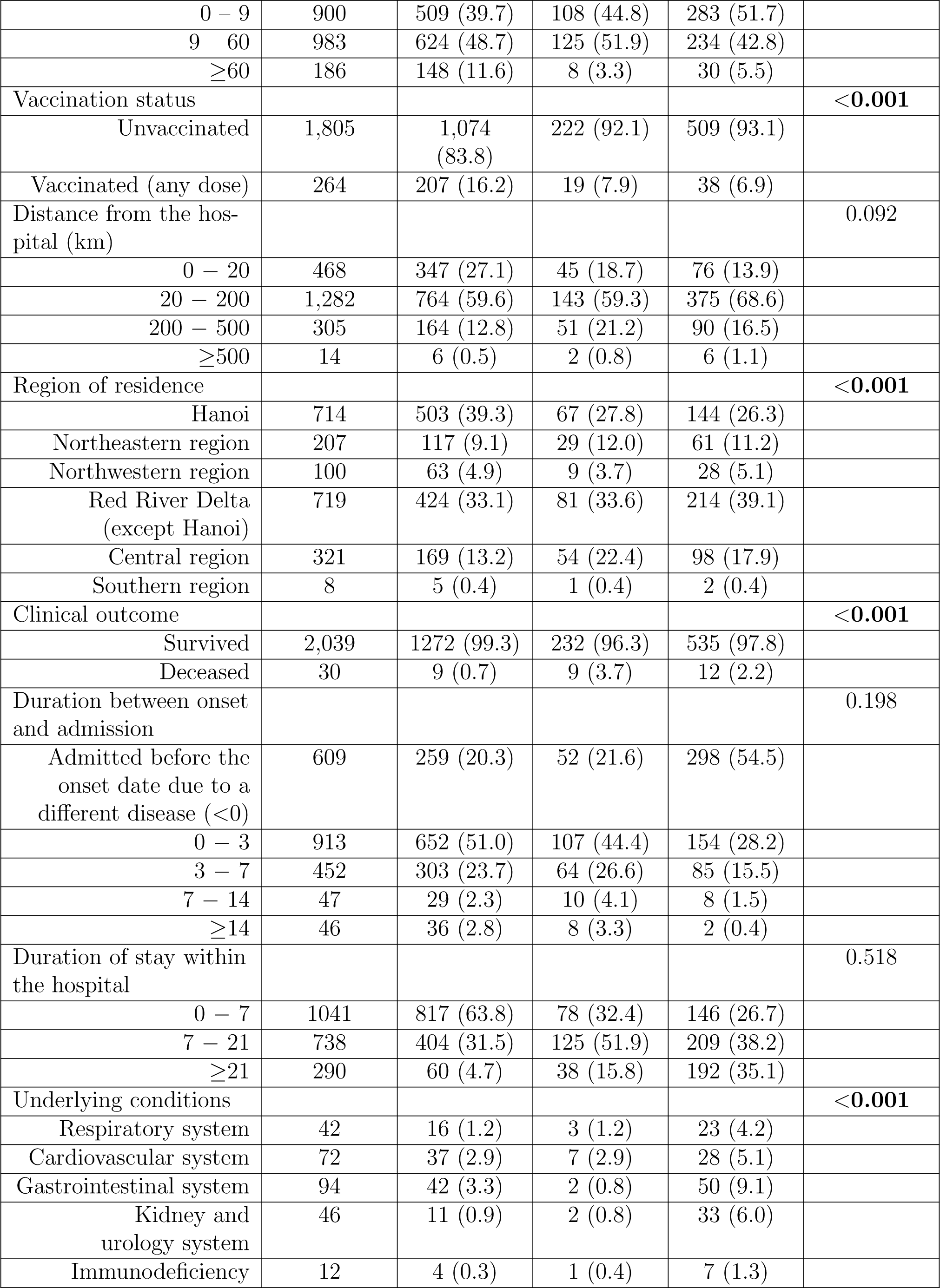
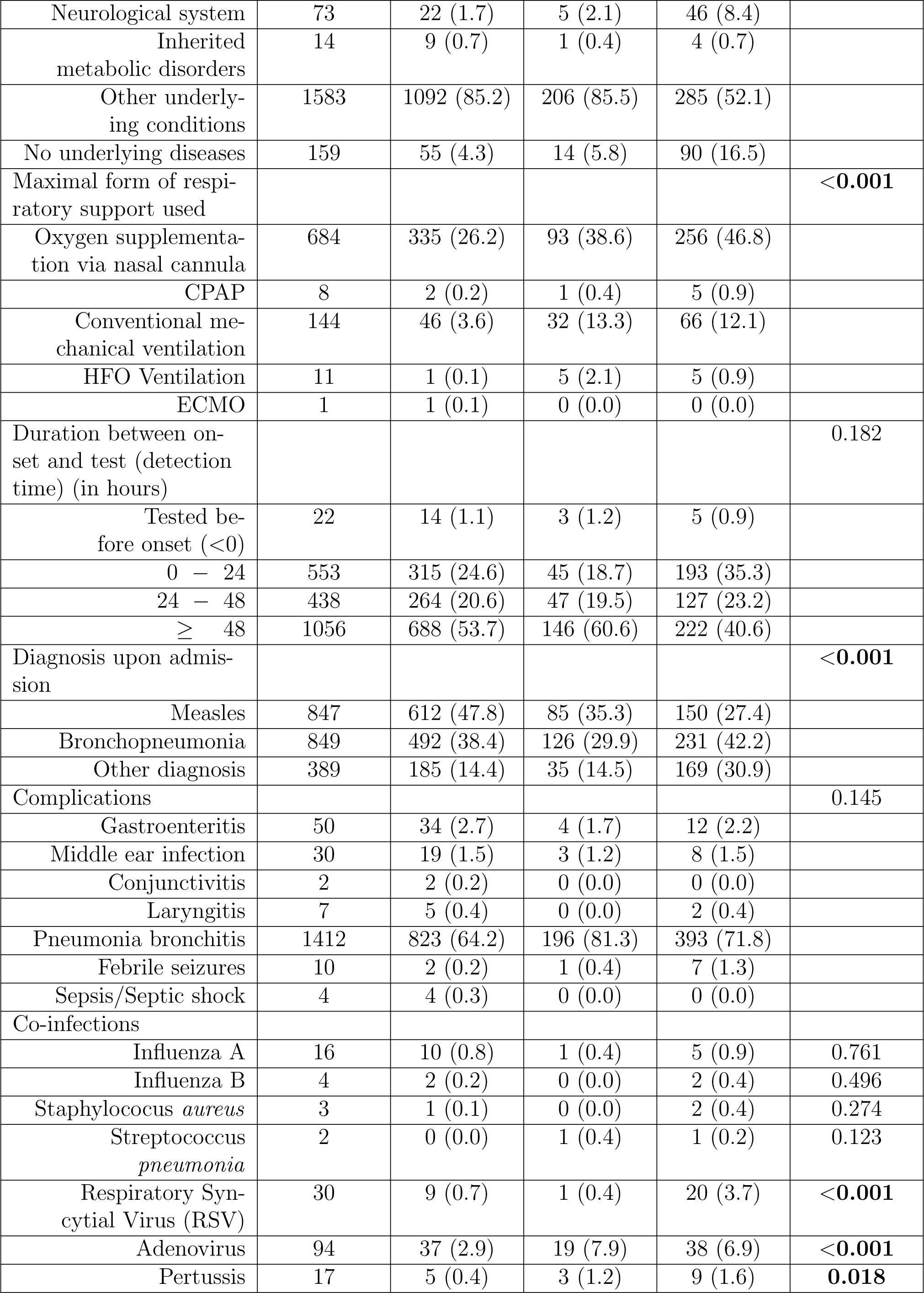
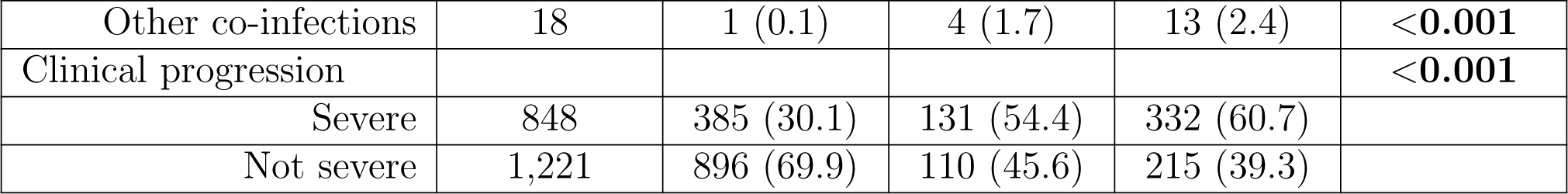
Demographic and clinical characteristics of hospitalized patients with measles by place of exposure Footnote: CPAP: Continuous Positive Pressure Ventilation, HFOV: High Frequency Oscillatory Ventilation, ECMO: Extracorporeal Membrane Oxygenation.

Table 3 depicts the relationships between socio-demographic, clinical characteristics and severity (turning severe). In the univariate analysis, the following factors were significantly associated with the likelihood of becoming severe: admission in 2018 and 2019, age 9 months to 5 years and more than 5 years, vaccination at least 1 dose, hospital transmission, living between 20 and 500 kilometres near VNCH, living in the Northeast, Red River Delta (except Hanoi), or Central Vietnam, mortality, being admitted to the hospital before the onset, hospitalized for more than a week, being tested more than 48 hours after the onset, having underlying diseases on the cardiovascular system, gastrointestinal system, kidney and urology system, neurological system. Having other underlying conditions or no underlying diseases is also heavily linked with severity. Moreover, being diagnosed with measles, or bronchopneumonia, having gastroenteritis, middle ear infection, laryngitis, or febrile seizures, having respiratory syncytial virus (RSV), adenovirus, pertussis, or other co-infections are connected to the risk of severity. Children with the following risk factors had significantly higher/lower odds of severity in the multivariable analysis: admission in 2018 (adjusted OR [AOR] 12.92, 95% CI 7.05 *−* 24.84) and in 2019 (AOR 16.16, 95% CI 8.95 *−* 30.69) compared with 2017, age under 9 months (AOR 2.60, 95% CI 1.64 *−* 4.20) and age from 9 months to 5 years (AOR 1.75, 95% CI 1.12 *−* 2.81) compared with less than 9 months, unvaccinated (AOR 1.56, 95% CI 1.09 *−* 2.27) compared with vaccinate, hospital transmission (AOR 1.73, 95% CI 1.38 *−* 2.17) compared to transmission in community, residing 20 to 200 kilometres (AOR 1.46, 95% CI 1.12 *−* 1.93) and 200 to 500 kilometres from VNCH (AOR 1.75, 95% CI 1.21 *−* 2.53) compared with less than 20 kilometres, hospitalized 7 to 21 days (AOR 3.50, 95% CI 2.79 *−* 4.40) and more than 21 days (AOR 14.20, 95% CI 9.15 *−* 22.04) compared with less than 7 days, having underlying condition in gastrointestinal system (AOR 6.74, 95% CI 1.65 *−* 28.87). Year of admission, age group, vaccination status, place of exposure, distance from VNCH, region of residence, clinical outcome, duration between onset and admission, duration of stay within VNCH had overall significance (*p <* 0.001). The association between duration between onset and test and severity had overall significance (*p* = 0.017 by likelihood ratio test), but not within an individual category. Factors no longer statistically significant in the multivariate analysis: having medical conditions in cardiovascular system, kidney and urology system, or neurological system, absence of underlying medical or other medical conditions, being tested 48 hours after onset and being diagnosed with measles, or bronchopneumonia, having RSV, adenovirus, pertussis or other co-infections. Sex, an unknown number of vaccinations, having inherited metabolic disorders, or the presence of an unknown place of exposure were not significantly associated with mortality in the uni- or multivariable analyses.

**Table 3:** Factors associated with the risk of severity among hospitalized patients with measles.

| Characteristics<br>(*N* = 2,072) | Total<br>(by<br>cate-<br>gory) | N (%) by<br>severity of<br>progression |  | Crude OR | p-value<br>(OR) | aOR | p-value<br>(aOR) |
| --- | --- | --- | --- | --- | --- | --- | --- |
|  |  | Severe | Not<br>severe |  |  |  |  |
| Number of patients | 2,072 | 849<br>(41.0) | 1,223<br>(59.0) |  |  |  |  |
| Year of admission |  |  |  |  | <0.001 |  | <0.001 |
| 2017 | 132 | 23<br>(2.7) | 109<br>(8.9) | Ref |  | Ref |  |
| 2018 | 621 | 246<br>(29.0) | 375<br>(30.7) | 3.06<br>(1.94 – 5.02) | <0.001 | 12.92<br>(7.05 – 24.84) | <0.001 |
| 2019 | 1,319 | 580<br>(68.3) | 739<br>(60.4) | 3.66<br>(2.36 – 5.92) | <0.001 | 16.16<br>(8.95 – 30.69) | <0.001 |
| Sex |  |  |  |  |  |  |  |
| Male | 1,297 | 542<br>(63.8) | 755<br>(61.7) | Ref |  | Ref |  |
| Female | 775 | 307<br>(36.2) | 468<br>(38.3) | 0.91<br>(0.76 – 1.09) | 0.331 | 1.06<br>(0.85 – 1.31) | 0.603 |
| Age in months<br>(mean, SD) |  | 13.7<br>(20.6) | 24.2<br>(32.9) |  |  |  |  |
| Age group (in months) |  |  |  |  | <0.001 |  | <0.001 |
| 0 – 9 | 902 | 450<br>(53) | 452<br>(37) | 4.10<br>(2.82 – 6.11) | <0.001 | 2.60<br>(1.64 – 4.20) | <0.001 |
| 9 – 60 | 984 | 363<br>(42.8) | 621<br>(50.8) | 2.41<br>(1.75 – 3.59) | <0.001 | 1.75<br>(1.12 – 2.81) | 0.014 |
| ≥60 | 186 | 36<br>(4.2) | 150<br>(12.3) | Ref |  | Ref |  |
| Vaccination status |  |  |  |  |  |  |  |
| Vaccinated (any dose) | 264 | 62<br>(7.3) | 202<br>(16.5) | Ref |  | Ref |  |
| Unvaccinated | 1,808 | 787<br>(92.7) | 1021<br>(83.5) | 2.50<br>(1.86 – 3.39) | <0.001 | 1.56<br>(1.09 – 2.27) | 0.015 |
| Place of exposure |  |  |  |  | <0.001 |  | <0.001 |
| Community exposed | 1,281 | 385<br>(45.3) | 896<br>(73.3) | Ref |  | Ref |  |
| Healthcare fa-<br>cility (including<br>VNCH and other) | 788 | 463<br>(54.5) | 325<br>(26.6) | 3.31<br>(2.75 – 3.98) | <0.001 | 1.73<br>(1.38 – 2.17) | <0.001 |
| Unknown | 3 | 1<br>(0.1) | 2<br>(0.2) | 1.40<br>(0.13 – 10.52) | 0.751 | 1.98<br>(0.14 – 29.36) | 0.580 |
| Distance from the hos-<br>pital (km) |  |  |  |  | <0.001 |  | <0.001 |
| 0 – 20 | 468 | 125<br>(14.7) | 343<br>(28.0) | Ref |  | Ref |  |
| 20 – 200 | 1285 | 567<br>(66.8) | 718<br>(58.7) | 2.16<br>(1.72 – 2.73) | <**0.001** | 1.46<br>(1.11 – 1.93) | **0.005** |
| 200 – 500 | 305 | 153<br>(18.0) | 152<br>(12.4) | 2.75<br>(2.04 – 3.74) | <**0.001** | 1.75<br>(1.21 – 2.53) | **0.003** |
| ≥500 | 14 | 4<br>(0.5) | 10<br>(0.8) | 1.17<br>(0.34 – 3.42) | 0.783 | 0.26<br>(0.05 – 1.06) | 0.061 |
| Region of residence |  |  |  |  | <**0.001** |  |  |
| Hanoi | 714 | 239<br>(28.2) | 475<br>(38.8) | Ref |  | N/A |  |
| Northeastern region | 207 | 98<br>(11.5) | 109<br>(8.9) | 1.79<br>(1.30 – 2.44) | <**0.001** | N/A |  |
| Northwestern region | 100 | 49<br>(5.8) | 51<br>(4.2) | 1.91<br>(1.25 – 2.91) | **0.002** | N/A |  |
| Red River Delta<br>(except Hanoi) | 722 | 306<br>(36.0) | 416<br>(34.0) | 1.46<br>(1.18 – 1.81) | <**0.001** | N/A |  |
| Central region | 321 | 155<br>(18.3) | 166<br>(13.6) | 1.85<br>(1.42 – 2.42) | <**0.001** | N/A |  |
| Southern region | 8 | 2<br>(0.2) | 6<br>(0.5) | 0.76<br>(0.14 – 3.01) | 0.714 | N/A |  |
| Clinical outcome |  |  |  |  | <**0.001** |  |  |
| Survived | 2042 | 819<br>(96.5) | 1223<br>(100.0) | Ref |  | N/A |  |
| Deceased | 30 | 30<br>(3.5) | 0<br>(0.0) | 91.07<br>(12.83 – 11500) | <**0.001** | N/A |  |
| Duration between onset<br>and admission |  |  |  |  | <**0.001** |  |  |
| Admitted before the<br>onset date due to a<br>different disease (<0) | 611 | 339<br>(39.9) | 272<br>(22.2) | 2.33<br>(1.89 – 2.88) | <**0.001** | N/A |  |
| 0 – 3 | 914 | 318<br>(37.5) | 596<br>(48.7) | Ref |  | N/A |  |
| 3 – 7 | 452 | 156<br>(18.4) | 296<br>(24.2) | 0.99<br>(0.78 – 1.25) | 0.924 | N/A |  |
| 7 – 14 | 47 | 22<br>(2.6) | 25<br>(2.0) | 1.65<br>(0.92 – 2.96) | 0.094 | N/A |  |
| ≥14 | 46 | 13<br>(1.5) | 33<br>(2.7) | 0.75<br>(0.38 – 1.41) | 0.385 | N/A |  |
| Unknown | 2 | 1<br>(0.1) | 1<br>(0.1) | 1.87<br>(0.15 – 23.13) | 0.591 | N/A |  |
| Duration of stay within<br>the hospital |  |  |  |  | <**0.001** |  | <**0.001** |
| 0 – 7 | 1042 | 226<br>(26.6) | 816<br>(66.7) | Ref |  | Ref |  |
| 7 – 21 | 739 | 391<br>(46.1) | 348<br>(28.5) | 4.05<br>(3.30 – 4.98) | <**0.001** | 3.50<br>(2.79 – 4.40) | <**0.001** |
| $\geq 21$ | 291 | 232<br>(27.3) | 59<br>(4.8) | 14.09<br>(10.29 – 19.55) | **<0.001** | 14.20<br>(9.15 – 22.57) | **<0.001** |
| Underlying conditions |  |  |  |  |  |  |  |
| Respiratory system | 42 | 23<br>(2.7) | 19<br>(1.6) | 1.76<br>(0.96 – 3.25) | 0.068 | 1.66<br>(0.38 – 7.67) | 0.503 |
| Cardiovascular system | 72 | 45<br>(5.3) | 27<br>(2.2) | 2.46<br>(1.53 – 4.03) | **<0.001** | 3.86<br>(0.90 – 17.61) | 0.070 |
| Gastrointestinal system | 94 | 73<br>(8.6) | 21<br>(1.7) | 5.29<br>(3.31 – 8.83) | **<0.001** | 6.74<br>(1.65 – 28.87) | **0.008** |
| Kidney and urology system | 46 | 20<br>(2.4) | 26<br>(2.1) | 1.12<br>(0.62 – 1.99) | **<0.001** | 0.95<br>(0.28 – 3.123) | 0.938 |
| Immunodeficiency | 12 | 6<br>(0.7) | 6<br>(0.5) | 1.44<br>(0.47 – 4.41) | 0.511 | 1.72<br>(0.26 – 11.74) | 0.575 |
| Neurological system | 73 | 44<br>(5.2) | 29<br>(2.4) | 2.24<br>(1.40 – 3.63) | **<0.001** | 3.31<br>(0.91 – 12.37) | 0.069 |
| Inherited metabolic disorders | 15 | 7<br>(0.8) | 8<br>(0.7) | 1.27<br>(0.46 – 3.44) | 0.633 | 1.64<br>(0.30 – 8.94) | 0.563 |
| Other underlying conditions | 1585 | 90<br>(10.6) | 69<br>(5.6) | 1.98<br>(1.43 – 2.75) | **<0.001** | 2.50<br>(0.61 – 10.56) | 0.202 |
| No underlying diseases | 159 | 554<br>(65.3) | 1031<br>(84.3) | 0.35<br>(0.28 – 0.43) | **<0.001** | 1.50<br>(0.38 – 6.13) | 0.560 |
| Maximal form of respiratory support used |  |  |  |  |  |  |  |
| Oxygen supplementation via nasal cannula | 685 | 685<br>(80.7) | 0<br>(0.0) | N/A | N/A | N/A |  |
| CPAP | 8 | 8<br>(0.9) | 0<br>(0.0) | N/A | N/A | N/A |  |
| Conventional mechanical ventilation | 144 | 144<br>(17.0) | 0<br>(0.0) | N/A | N/A | N/A |  |
| FHO Ventilation | 11 | 11<br>(1.3) | 0<br>(0.0) | N/A | N/A | N/A |  |
| ECMO | 1 | 1<br>(0.1) | 0<br>(0.0) | N/A | N/A | N/A |  |
| Duration between onset and test (detection time) (in hours) |  |  |  |  | **0.017** |  | 0.223 |
| Tested before onset (<0) | 23 | 8<br>(0.9) | 15<br>(1.2) | 0.72<br>(0.29 – 1.65) | 0.439 | 1.32<br>(0.45 – 3.61) | 0.605 |
| 0 – 24 | 554 | 240<br>(28.3) | 314<br>(25.7) | Ref |  | Ref |  |
| 24 – 48 | 438 | 201<br>(23.7) | 237<br>(19.4) | 1.11<br>(0.86 – 1.43) | 0.419 | 1.33<br>(0.98 – 1.82) | 0.069 |
| $\geq 48$ | 1057 | 400<br>(47.1) | 657<br>(53.7) | 0.80<br>(0.65 – 0.98) | **0.033** | 1.28<br>(0.99 – 1.67) | 0.063 |
| Diagnosis upon admission |  |  |  |  |  |  |  |
| Measles | 848 | 272<br>(32.0) | 576<br>(47.1) | 0.53<br>(0.44 – 0.64) | **<0.001** | 0.77<br>(0.19 – 3.26) | 0.721 |
| Bronchopneumonia | 849 | 409<br>(48.2) | 440<br>(36.0) | 1.36<br>(1.14 – 1.63) | **<0.001** | 1.06<br>(0.26 – 4.48) | 0.940 |
| Other diagnosis | 391 | 177<br>(20.8) | 214<br>(17.5) | 1.24<br>(0.99 – 1.55) | 0.056 | 0.49<br>(0.14 – 2.58) | 0.480 |
| Complications |  |  |  |  |  |  |  |
| Gastroenteritis | 51 | 14<br>(1.6) | 37<br>(3.0) | 0.55<br>(0.29 – 0.99) | **0.047** | N/A |  |
| Middle ear infection | 30 | 4<br>(0.5) | 26<br>(2.1) | 0.24<br>(0.08 – 0.60) | **0.001** | N/A |  |
| Conjunctivitis | 2 | 1<br>(0.1) | 1<br>(0.1) | 1.44<br>(0.12 – 17.76) | 0.753 | N/A |  |
| Laryngitis | 7 | 0<br>(0.0) | 7<br>(0.6) | 0.09<br>(0.00 – 0.78) | **0.024** | N/A |  |
| Pneumonia bronchitis | 1413 | 721<br>(84.9) | 692<br>(56.6) | 4.31<br>(3.47 – 5.38) | **<0.001** | N/A |  |
| Febrile seizures | 10 | 9<br>(1.1) | 1<br>(0.1) | 9.21<br>(2.12 – 86.00) | **0.001** | N/A |  |
| Sepsis/Septic shock | 4 | 2<br>(0.2) | 2<br>(0.2) | 1.44<br>(0.22 – 9.34) | 0.684 | N/A |  |
| Co-infections |  |  |  |  |  |  |  |
| Influenza A | 16 | 8<br>(0.9) | 8<br>(0.7) | 1.44<br>(0.55 – 3.82) | 0.452 | N/A |  |
| Influenza B | 4 | 3<br>(0.4) | 1<br>(0.1) | 3.37<br>(0.55 – 34.81) | 0.189 | N/A |  |
| *Staphylococcus aureus* | 3 | 1<br>(0.1) | 2<br>(0.2) | 0.86<br>(0.08 – 6.50) | 0.887 | N/A |  |
| *Streptococcus pneumoniae* | 2 | 2<br>(0.2) | 0<br>(0.0) | 7.22<br>(0.59 – 996) | 0.13 | N/A |  |
| Respiratory Syncytical Virus (RSV) | 30 | 18<br>(2.1) | 12<br>(1.0) | 2.16<br>(1.06 – 4.55) | **0.035** | 0.76<br>(0.32 – 1.86) | 0.541 |
| Adenovirus | 95 | 73<br>(8.6) | 22<br>(1.8) | 5.05<br>(3.18 – 8.35) | **<0.001** | 1.71<br>(0.97 – 3.11) | 0.212 |
| Pertussis | 17 | 12<br>(1.4) | 5<br>(0.4) | 3.31<br>(1.26 – 9.88) | **0.014** | 0.85<br>(0.26 – 2.99) | 0.788 |
| Other co-infections | 18 | 15<br>(1.8) | 3<br>(0.2) | 6.48<br>(2.25 – 24.88) | **<0.001** | 2.51<br>(0.62 – 13.95) | 0.212 |

Table 4 illustrates the relationships between socio-demographic and clinical factors and mortality. In the univariate analysis, the following factors were significantly associated with fatality: age 9 months to 5 years, community transmission, living 20 to 500 kilometres from VNCH, living in the Northeast, Northwest or Central Vietnam, being admitted to the hospital before the onset or after 2 weeks, hospitalized for more than a week, having no underlying diseases or other underlying con-ditions, requiring a form of respiratory support (oxygen supplementation via nasal cannula, CPAP, conventional mechanical ventilation, HFO ventilation). Furthermore, having adenovirus or other healthcare-related infections, being severe, having sepsis/septic shock sequelae, and being diagnosed with measles, bronchopneumonia, or other diseases on admission are all substantially linked with death. Children with the following risk factors had significantly higher/lower odds of mortality in the multivariable analysis: age under 9 months (adjusted OR [AOR] 0.21, 95% CI 0.07 *−* 0.70) and age from 9 months to 5 years (AOR 0.19, 95% CI 0.06 *−* 0.60) compared with less than 9 months, residing 20 to 200 kilometres from VNCH (AOR 4.09, 95% CI 1.12 *−* 23.19), having adenovirus (AOR 8.29, 95% CI 3.31*−*20.45) or other hospital acquired infections (AOR 7.20, 95% CI 1.14*−*40.39). The association between place of exposure and mortality had overall significance (*p* = 0.001 by likelihood ratio test), but not within an individual category. The duration between onset and admission, as well as duration of stay within the hospital, had overall significance (*p* = 0.015 and *p <* 0.001, respectively). Factors such as age less than 9 months, hospital transmission, hospitalization for more than one week, absence of underlying medical or other medical conditions, living 20 to 200 km away from VNCH and being diagnosed with measles, bronchopneumonia, or others on admission in the univariable analysis were no longer statistically significant in the multivariate analysis. Sex, admission year, having bronchopneumonia or the presence of an unknown place of exposure were not significantly associated with mortality in the uni- or multivariable analyses.

**Table 4:** Association with mortality among hospitalized patients.

| Characteristics (N = 2,072) | N (%) by final condition |  | Crude OR (95% Confidence Interval) | p-value (OR) | aOR (95% CI) | p-value (aOR) |
| --- | --- | --- | --- | --- | --- | --- |
|  | Deceased | Discharged |  |  |  |  |
| Number of patients | 30 (1.4) | 2,042 (98.6) |  |  |  |  |
| Year of admission |  |  |  | 0.485 |  |  |
| 2017 | 2 (6.7) | 130 (6.4) | Ref |  | Ref |  |
| 2018 | 6 (20.0) | 615 (30.1) | 0.55 (0.13 – 3.02) | 0.449 | 0.79 (0.16 – 5.23) | 0.791 |
| 2019 | 22 (73.3) | 1,297 (63.5) | 0.90 (0.28 – 4.52) | 0.884 | 1.43 (0.35 – 8.68) | 0.645 |
| Sex |  |  |  |  |  |  |
| Male | 18 (60.0) | 1,279 (62.6) | Ref |  | Ref |  |
| Female | 12 (40.0) | 763 (37.4) | 1.13 (0.53 – 2.31) | 0.737 | 1.47 (0.67 – 3.16) | 0.328 |
| Age in months (mean, SD) | 32.6 (43.1) | 19.7 (28.7) |  |  |  |  |
| Age group (in months) |  |  |  | 0.131 |  |  |
| 0 – 9 | 12 (40.0) | 890 (43.6) | 0.39 (0.15 – 1.09) | 0.07 | 0.21 (0.07 – 0.70) | **0.012** |
| 9 – 60 | 12 (40.0) | 972 (47.6) | 0.36 (0.14 – 0.99) | **0.049** | 0.19 (0.06 – 0.60) | **0.006** |
| ≥60 | 6 (20.0) | 180 (8.8) | Ref |  | Ref |  |
| Vaccination status |  |  |  | 0.824 |  |  |
| Vaccinated (any dose) | 4<br>(13.3) | 260<br>(12.7) | Ref |  | Ref |  |
| Unvaccinated | 26<br>(86.7) | 1782<br>(87.3) | 0.86<br>(0.35 – 2.72) | 0.774 | 0.50<br>(0.17 – 1.73) | 0.252 |
| Place of exposure |  |  |  | **0.001** |  |  |
| Community exposed | 385<br>(45.3) | 896<br>(73.3) | Ref |  | Ref |  |
| Health care facility (including VNCH and other) | 463<br>(54.5) | 325<br>(26.6) | 3.75<br>(1.79 – 8.49) | **<0.001** | 1.92<br>(0.82 – 4.72) | 0.132 |
| Unknown | 1<br>(0.1) | 2<br>(0.2) | 19.14<br>(0.14–218.87) | 0.116 | 6.72<br>(0.04–206.20) | 0.356 |
| Distance from the hospital (km) |  |  |  | 0.145 |  |  |
| 0 – 20 | 2<br>(6.7) | 466<br>(22.8) | Ref |  | Ref |  |
| 20 – 200 | 22<br>(73.3) | 1263<br>(61.9) | 3.32<br>(1.07 – 16.50) | **0.036** | 4.09<br>(1.12 – 23.20) | **0.031** |
| 200 – 500 | 6<br>(20.0) | 299<br>(14.6) | 4.04<br>(1.02 – 22.14) | **0.045** | 3.86<br>(0.82 – 24.48) | 0.088 |
| ≥500 | 0<br>(0.0) | 14<br>(0.7) | 6.43<br>(0.04 – 83.93) | 0.332 | 5.10<br>(0.03–118.76) | 0.433 |
| Region of residence |  |  |  | 0.077 |  |  |
| Hanoi | 4<br>(13.3) | 710<br>(34.8) | Ref |  | N/A |  |
| Northeastern region | 5<br>(16.7) | 202<br>(9.9) | 4.28<br>(1.20 – 16.08) | **0.025** | N/A |  |
| Northwestern region | 3<br>(10.0) | 97<br>(4.8) | 5.66<br>(1.25 – 23.61) | **0.026** | N/A |  |
| Red River Delta (except Hanoi) | 11<br>(36.7) | 711<br>(34.8) | 2.55<br>(0.90 – 8.57) | 0.077 | N/A |  |
| Central region | 7<br>(23.3) | 314<br>(15.4) | 3.76<br>(1.19 – 13.38) | **0.024** | N/A |  |
| Southern region | 0<br>(0.0) | 8<br>(0.4) | 9.28<br>(0.06 – 98.77) | 0.259 | N/A |  |
| Duration between onset and admission |  |  |  | **0.015** |  |  |
| Admitted before the onset date due to a different disease (<0) | 16<br>(53.3) | 595<br>(29.1) | 3.35<br>(1.45 – 8.49) | **0.004** | N/A |  |
| 0 – 3 | 7<br>(23.3) | 907<br>(44.4) | Ref |  | N/A |  |
| 3 – 7 | 4<br>(13.3) | 448<br>(21.9) | 1.21<br>(0.34 – 3.83) | 0.748 | N/A |  |
| 7 – 14 | 1<br>(3.3) | 46<br>(2.3) | 3.90<br>(0.40 – 18.40) | 0.195 | N/A |  |
| $\geq 14$ | 2<br>(6.7) | 44<br>(2.2) | 6.79<br>(1.24 – 26.36) | **0.03** | N/A | |
| Unknown | 0<br>(0.0) | 2<br>(0.1) | 24.20<br>(0.16 – 333.87) | 0.146 | N/A |  |
| Duration of stay within<br>the hospital |  |  |  | **<0.001** |  |  |
| 0 – 7 | 5<br>(16.7) | 1037<br>(50.8) | Ref |  | Ref |  |
| 7 – 21 | 13<br>(43.3) | 726<br>(35.6) | 3.50<br>(1.35 – 10.39) | **<0.001** | 2.14<br>(0.78 – 6.67) | 0.144 |
| $\geq 21$ | 12<br>(40.0) | 279<br>(13.7) | 8.43<br>(3.20 – 25.32) | **<0.001** | 1.60<br>(0.44 – 6.02) | 0.468 |
| Underlying conditions |  |  |  |  |  |  |
| Respiratory system | 1<br>(3.3) | 41<br>(2.0) | 2.45<br>(0.27 – 9.68) | 0.35 | 0.60<br>(0.00 – 62.94) | 0.871 |
| Cardiovascular system | 1<br>(3.3) | 71<br>(3.5) | 1.40<br>(0.15 – 5.44) | 0.703 | 0.15<br>(0.00 – 7.17) | 0.452 |
| Gastrointestinal system | 2<br>(6.7) | 92<br>(4.5) | 1.84<br>(0.37 – 5.72) | 0.395 | 0.54<br>(0.00 – 47.32) | 0.834 |
| Kidney and<br>urology system | 0<br>(0.0) | 46<br>(2.3) | 0.70<br>(0.00 – 5.13) | 0.795 | 0.10<br>(0.00 – 2.25) | 0.157 |
| Immunodeficiency | 1<br>(3.3) | 11<br>(0.5) | 8.98<br>(0.95 – 39.64) | 0.053 | 1.48<br>(0.00 – 200.11) | 0.900 |
| Neurological system | 2<br>(6.7) | 71<br>(3.5) | 2.41<br>(0.48 – 7.52) | 0.239 | 0.81<br>(0.00 – 59.76) | 0.941 |
| Inherited<br>metabolic disorders | 0<br>(0.0) | 15<br>(0.7) | 2.14<br>(0.01 – 16.59) | 0.637 | 0.65<br>(0.00 – 105.03) | 0.896 |
| Other underly-<br>ing conditions | 8<br>(26.7) | 151<br>(7.4) | 4.71<br>(1.99 – 10.20) | **<0.001** | 0.78<br>(0.00 – 71.50) | 0.933 |
| No underlying diseases | 15<br>(50.0) | 1570<br>(76.9) | 0.30<br>(0.14 – 0.61) | **0.001** | 0.25<br>(0.00 – 23.15) | 0.630 |
| Maximal form of respi-<br>ratory support used |  |  |  |  |  |  |
| Oxygen supplementa-<br>tion via nasal cannula | 2<br>(6.8) | 683<br>(33.4) | 7.19<br>(3.06 – 20.29) | **<0.001** | N/A |  |
| CPAP | 0<br>(0.0) | 8<br>(0.4) | 7.29<br>(2.24 – 18.92) | **0.002** | N/A |  |
| Conventional me-<br>chanical ventilation | 21<br>(70.0) | 123<br>(6.0) | 169.84<br>(55.25 – 840.85) | **<0.001** | N/A |  |
| HFO Ventilation | 7<br>(23.3) | 4<br>(0.2) | 118.23<br>(36.86 – 403.66) | **<0.001** | N/A |  |
| ECMO | 0<br>(0.0) | 1<br>(0.0) | 22.31<br>(0.15 – 426.93) | 0.156 | N/A |  |
| Severity |  |  |  | **<0.001** |  |  |
| Severe | 30<br>(100.0) | 819<br>(40.1) | 91.07<br>(12.83 – 11541.95) | <**0.001** | N/A |  |
| Not severe | 0<br>(0.0) | 1223<br>(59.9) | Ref |  | N/A |  |
| Duration between onset and test (detection time) (in hours) |  |  |  | 0.26 |  |  |
| Tested before onset (<0) | 1<br>(3.3) | 15<br>(0.7) | 3.88<br>(0.40 – 18.30) | 0.197 | N/A |  |
| 0 – 24 | 10<br>(33.3) | 421<br>(20.6) | Ref |  | N/A |  |
| 24 – 48 | 4<br>(13.3) | 427<br>(20.9) | 0.42<br>(0.12 – 1.21) | 0.113 | N/A |  |
| ≥48 | 15<br>(50.0) | 1177<br>(57.6) | 0.52<br>(0.24 – 1.20) | 0.123 | N/A |  |
| Unknown | 0<br>(0.0) | 2<br>(0.1) | 8.02<br>(0.05–107.16) | 0.29 | N/A |  |
| Diagnosis upon admission |  |  |  |  |  |  |
| Measles | 6<br>(20.0) | 842<br>(41.2) | 0.37<br>(0.14 – 0.85) | **0.017** | 3.41<br>(0.31 – 21.90) | 0.284 |
| Bronchopneumonia | 16<br>(53.3) | 833<br>(40.8) | 1.65<br>(0.81 – 3.40) | 0.017 | 8.45<br>(0.68 – 67.82) | 0.090 |
| Other diagnosis | 10<br>(33.3) | 381<br>(18.7) | 2.23<br>(1.01 – 4.63) | **0.046** | 7.33<br>(0.48 – 76.63) | 0.143 |
| Complications |  |  |  |  |  |  |
| Gastroenteritis | 0<br>(0.0) | 51<br>(2.5) | 0.63<br>(0.00 – 4.61) | 0.731 | N/A |  |
| Middle ear infection | 0<br>(0.0) | 30<br>(1.5) | 1.08<br>(0.00 – 8.01) | 0.956 | N/A |  |
| Conjunctivitis | 0<br>(0.0) | 2<br>(0.1) | 13.38<br>(0.09–169.03) | 0.208 | N/A |  |
| Laryngitis | 0<br>(0.0) | 7<br>(0.3) | 4.44<br>(0.03 – 37.94) | 0.405 | N/A |  |
| Pneumonia bronchitis | 25<br>(83.3) | 1388<br>(68.0) | 2.18<br>(0.93 – 6.16) | 0.074 | N/A |  |
| Febrile seizures | 0<br>(0.0) | 10<br>(0.5) | 3.17<br>(0.02 – 25.64) | 0.499 | N/A |  |
| Sepsis/Septic shock | 1<br>(3.3) | 3<br>(0.1) | 29.63<br>(2.81–186.83) | **0.009** | N/A |  |
| Co-infections |  |  |  |  |  |  |
| Influenza A | 0<br>(0.0) | 16<br>(0.8) | 2.01<br>(0.01 – 15.50) | 0.662 | N/A |  |
| Influenza B | 0<br>(0.0) | 4<br>(0.2) | 7.42<br>(0.05 – 71.98) | 0.295 | N/A |  |
| Staphylococcus *aureus* | 0<br>(0.0) | 3<br>(0.1) | 9.55<br>(0.07–101.65) | 0.254 | N/A |  |
| Streptococcus<br>*pneumonia* | 0<br>(0.0) | 2<br>(0.1) | 3.38<br>(0.09–169.03) | 0.208 | N/A |  |
| Respiratory Syn-<br>cytical Virus (RSV) | 1<br>(3.3) | 29<br>(1.4) | 3.47<br>(0.38 – 13.94) | 0.217 | N/A |  |
| Adenovirus | 11<br>(36.7) | 84<br>(4.1) | 13.66<br>(6.21 – 28.85) | <**0.001** | 8.29<br>(3.31 – 20.45) | <**0.001** |
| Pertussis | 0<br>(0.0) | 17<br>(0.8) | 1.89<br>(0.01 – 14.53) | 0.686 | N/A |  |
| Other co-infections | 3<br>(10.0) | 15<br>(0.7) | 16.64<br>(4.16 – 51.24) | <**0.001** | 7.20<br>(1.14 – 40.39) | **0.037** |

## 4. Dicussions

In this study, we present the epidemiological and clinical characteristics of more than 2,000 children hospitalized with measles in Hanoi, the vast majority during Vietnam’s 2017-19 epidemic. To the best of our knowledge, this study is one of the few that provides data on the number of severe cases. According to the caregiver report or immunization card, 1,808 (87.3%) had not received any MCV. More than half of the individuals in our study reported the community as the likely place of infection exposure, suggesting secondary cases. 69.3% of patients live in the Red River Delta, with 34.5% living in Hanoi, Vietnam’s second most densely populated region, with a population of 2209, 2239, and 2410 per square kilometre in 2017, 2018, and 2019 respectively [16, 17, 18]. Crowded environments and high concentrations of people have known risk factors for disease occurrence and propagation, and Hanoi has been cited as a hotspot of prior epidemics in the country [19, 20]. During the epidemic, children admitted from further distances are most likely referrals from other hospitals due to complications. Despite the high number of hospital admissions and the severity rate, the mortality rate of 1.4% discovered in this study of patients hospitalized with measles is relatively low lower than the global norm. A systematic review and modelling analysis, encompassing various studies, reported an overall pooled mean case fatality rate for measles of 2.2%, with an in-hospital mortality rate ranging from 0.9% to 6.0%, higher than the rate observed in this study [21]. The lower in-hospital mortality rate observed here may be attributed to several factors, including increased measles vaccination coverage, heightened public awareness and recognition, a higher rate of hospitalization, and prompt treatment. Additionally, the fact that the study was conducted at a national-level hospital, where patients with severe symptoms receive appropriate medical care, may have contributed to the lower mortality rate.

The median age of children was under 10 months. Children aged 9 months to 5 years accounted for 12 (60%) of the 30 deaths. To boost MCV1 coverage and reduce cases and deaths in this age range, routine immunization must be strengthened. The strategy is less obvious for the 40% of deaths in infants under the age of 9 months, before their scheduled MCV1. Measles vaccination at 4 to 5 months of age was beneficial in preventing serologically confirmed and definite clinical measles during an outbreak in Guinea-Bissau in 2003-2004 [22]. Vietnam recommends measles vaccination for children from 9 months and in late 2018, 4.2 million children aged 1 to 5 received measles vaccine as part of an additional measles-rubella immunization campaign [9, 23]. In the event of future epidemics, these measures must be swift and effective. The earlier timing of MCV in low-income settings where maternal antibody levels may be low requires further evaluation. Upon admission, 849 (41.0%) children have bronchopneumonia. Most children had complications, with bronchopneumonia, the most common measles complication, observed in two-thirds of cases (68.2%). This prevalence of bronchopneumonia was higher than that reported in Greece (46.2%), Israel (30.5%), China (33%), and Serbia (26.8%), although these studies included adolescent cases, which could have affected the aggregated results [24, 25, 26, 27]. Tu et al. reported that measles-associated bronchopneumonia was associated with younger age, which explains the higher rate of bronchopneumonia in our study [26]. There was no additional information on the pathogens causing pneumonia or gastroenteritis, or the nature of the neurological complications.

There were no statistically significant differences in the risk of severe complications or mortality between the two genders. The odds of death increased by more than three times for children living 20 to 200 kilometres away from the hospital and by more than four times for children living 200 to 500 kilometres away, likely reflecting the increment in travel time to VNCH and referrals from other healthcare institutions due to complications. Children with bronchopneumonia complications had a greater risk of death, as expected and consistent with earlier research [28, 29]. Compared to children who had received one or more doses of MCV, the odds of severity were more than three times greater in unvaccinated children. Vaccine-modified measles is characterized by a milder illness in children with preexisting protection following vaccination [30, 31]. Patients with community transmission have lower risk of severe forms of disease and mortality rates than patients exposed to other sources. The severity rate increased fivefold and the mortality rate increased more than tenfold with adenovirus infection.

It is not a question of if, but when the next measles epidemic will occur in Vietnam and other countries. Reported measles cases have reduced from the last six months of 2020 to 2022, likely due to COVID-19 social distancing measures, reduced domestic and international travel, disturbance in surveillance and reporting, and the phase of the measles epidemic cycle [7]. Furthermore, the Vietnam Ministry of Health listed measles on the list of group B infectious diseases requiring medical isolation as well as expanded vaccination for subjects aged 6-9 months, combined with other reasonable measures to control epidemics from hospitals, limiting the spread of measles in hospitals [32]. However, at the same time, the COVID-19 pandemic has significantly disrupted routine vaccination practices, with estimates of more than 90 million children missing measles vaccine doses by October 2020, and reports of a lost generation of children [33]. MCV1 coverage fell from an average of 85% during 2015-2019 to 84% in 2020 and 81% in 2021, the lowest coverage levels for MCV1 since 2008. The most significant declines in MCV1 coverage during 2019-2021 occurred in the South-East Asia Region (from 94% to 86%) and the Western Pacific Region (from 95% to 91%). MCV2 coverage grew from 63% to 71% between 2015 and 2019, suggesting second-dose introductions in many countries. However, MCV2 coverage remained stable thereafter (72% in 2020 and 71% in 2021), with only four additional countries deploying MCV2 during 2020-2021 [34]. Vaccination coverage in Vietnam declined further to 89% for MCV1 and 85% for MCV2 in 2021 [35]. When COVID-19 limitations are lifted, an increasing number of unvaccinated children who are susceptible to measles creates an environment for measles to resurface [36]. Repeated occurrences of measles outbreaks in Vietnam highlight the significance of re-evaluating measles virus dynamics and prevention and control strategies. Actions are required to reach unimmunised children through catch-up campaigns and to prepare for anticipated outbreaks. Governments must not lose sight of the Measles and Rubella Strategic Framework, 2021-2030 [37].

This study has certain limitations. The patients enrolled in this study are those that were admitted to our hospital, whose illness may be more severe and are not representative of all measles cases in Vietnam. Nonetheless, since patients come from various regions across the country, they still offer insights into the broader measles landscape in Vietnam. Detailed data were partially lacking on the number of MCVs and the place of exposure though the medical files were thoroughly checked, and the percentage of missing data was quite low for almost all variables. Nonetheless, we were able to analyze a large sample of over 2,000 measles cases with extensive clinical information and other socio-demographic characteristics using the measles suspected case inquiry form.

In conclusion, in this study, we report clinical and epidemiological characteristics from a significant measles outbreak that primarily affected unvaccinated children admitted to VNCH with significant number of deaths occurred among infants aged less than nine months. As MCV1 coverage in Vietnam declined to 89% in 2021, routine immunization needs to be strengthened and the earlier timing of MCV1 requires further evaluation to prevent further outbreaks.

## Data Availability

All data produced in the present study are available upon reasonable request to the authors.

